# A Custom Global Screening Array for Integrated Familial Hypercholesterolemia Detection and Polygenic Risk Assessment in a Multi-Ethnic New Zealand Population

**DOI:** 10.64898/2026.06.22.26355820

**Authors:** Alexandr Vikhorev, Maksim Struchalin, Xiaoyan Sun, Yalu Wen, Helen Wihongi, Patrick Gladding

## Abstract

**Background:** Cardiovascular disease (CVD) is the leading cause of mortality in New Zealand, with significant inequities affecting Māori and Pacific peoples. Familial hypercholesterolemia (FH) affects approximately 1 in 313 individuals globally, yet over 90% remain undiagnosed. Standard polygenic risk scores (PRS) derived from European cohorts may not be portable to diverse ancestries. We developed the Holo-Q Omniscan Waka Te Ira, a custom Illumina Global Screening Array (GSA) v3 enriched with FH mutations, CAD PRS markers, and network medicine-derived epistasis content.

**Methods:** Holo-Q Omniscan Waka Te Ira was developed as a customized version of the Illumina Global Screening Array v3, adding 43,437 SNPs targeting familial hypercholesterolaemia and coronary artery disease. Variants in the three primary FH genes were collected from a regional diagnostic laboratory list, published literature, ClinVar, and the Leiden Open Variation Database, resulting in 6,717 unique SNPs. Additional content included 14,005 pathogenic or likely pathogenic variants from cardiovascular, lipid-related genes and 12 pharmacogenes; and 5,845 probes supporting copy number variant detection. The array further incorporated 5,232 network medicine–derived coronary artery disease SNPs and 14,806 rare variants from a validated multi-ancestry polygenic score. To improve ancestral representation, 289 variants specific to New Zealand, European, Asian, and African populations, along with 118 variants specific to populations from Japan, Korea, Thailand, and Russia were added. Validation was performed using large-scale genotype and whole-genome sequencing datasets with polygenic score benchmarking. The completed design contained 47,027 SNPs overall, including 3,590 loci inherited from the GSA v3 backbone and 43,437 newly incorporated through custom content expansion.

**Results:** Approximately half of the newly added SNPs were observed in a large European-ancestry dataset, with high recovery for common polygenic score loci but low recovery for population-specific founder variants. The array captured 938 (84%) of all unique pathogenic or likely pathogenic familial hypercholesterolaemia variants catalogued in ClinVar at the time of design, representing a 26.4% expansion beyond the standard backbone array. Whole-genome sequencing validation identified additional carriers of rare high-impact variants present only in the custom content. The selected coronary artery disease polygenic score model achieved an adjusted area under the receiver operating characteristic curve of 0.786. Together, these results demonstrate enhanced monogenic detection, robust polygenic performance, and improved representation of ancestrally diverse populations within a single screening platform.

**Conclusion:** The Holo-Q Omniscan Waka Te Ira enhances detection of clinically relevant FH variants and provides robust PRS coverage. The low recovery of population-specific alleles in UK datasets underscores the necessity of this custom array for equitable genomic medicine in New Zealand’s multi-ethnic population.

## 1. Introduction

### 1.1 The Burden of Cardiovascular Disease in New Zealand

Cardiovascular disease (CVD) remains the leading cause of death and disability in New Zealand, accounting for over 30% of all deaths annually (1). Acute coronary syndrome (ACS) and inherited lipid disorders, particularly familial hypercholesterolemia (FH), are major contributors to this burden. Despite advances in treatment, many individuals at high risk remain undiagnosed or inadequately managed, resulting in preventable morbidity and mortality. Given the high burden of CVD in Māori and Pacific peoples, it is imperative that New Zealand implement genetic technologies to reduce CVD risk in an equitable manner.

### 1.2 Familial Hypercholesterolemia: Prevalence, Diagnosis, and Gaps

Familial hypercholesterolemia is the most common monogenic disorder, affecting approximately 1 in 313 individuals globally (2). In New Zealand, the prevalence is similar, but up to 90% of cases remain undiagnosed (3). FH is associated with lifelong elevated low-density lipoprotein cholesterol (LDL-C) and a markedly increased risk of premature atherosclerotic cardiovascular disease (ASCVD). Early detection and treatment with lipid-lowering therapies, such as statins, can reduce the risk of major adverse cardiovascular events (MACE) by up to 80% (4).

FH is primarily caused by mutations in three major genes: LDLR, APOB, and PCSK9. The LDLR gene, encoding the LDL receptor, is the most common cause; mutations reduce the receptor’s ability to clear LDL cholesterol from the blood. APOB gene mutations affect apolipoprotein B, which helps bind LDL to its receptor, leading to inefficient cholesterol clearance. Mutations in PCSK9 can increase its activity, resulting in reduced LDL receptor availability on liver cells and higher cholesterol levels.

Traditional diagnosis of FH relies on clinical criteria, lipid measurements, and family history. However, these methods have limited sensitivity and specificity, particularly in populations with high rates of metabolic syndrome or in older adults where secondary causes of hypercholesterolemia are common (4). Targeted genetic testing for pathogenic variants has improved diagnostic accuracy, but access remains limited and is often not tailored to the genetic diversity of New Zealand’s population (5,6).

### 1.3 Polygenic Risk and the Infinitesimal Model

Not all individuals with high cholesterol or premature heart disease have a single, identifiable genetic mutation. While monogenic FH is caused by rare, high-impact variants, a substantial proportion of patients with a clinical FH phenotype do not have a detectable mutation in these genes. Many of these cases are now understood to be due to the cumulative effect of multiple common genetic variants, each conferring a small increase in risk—a concept first formalised by Sir Ronald Fisher’s infinitesimal model in 1918 (7).

Polygenic risk scores (PRS) aggregate the effects of thousands of common variants, providing a quantitative measure of genetic predisposition to complex diseases like coronary artery disease (CAD) and hypercholesterolemia (8). In the context of FH, PRS offer several important advantages. First, they enable risk stratification among individuals with known monogenic FH, helping to identify those at highest risk for adverse cardiovascular outcomes (9). Second, PRS can identify individuals with “polygenic FH”—those who have high cholesterol due to the combined effect of many common variants rather than a single rare mutation (10). This is particularly valuable for patients who meet clinical criteria for FH but test negative for known monogenic mutations.

Moreover, PRS can help predict individuals who may carry a hidden or undetected mutation, as those with a low LDL PRS and a profile suggestive of FH may warrant further genetic investigation (11,12). Recent studies have shown that PRS can identify individuals at equivalent risk for heart disease as patients with FH, who would not be detected by traditional clinical or monogenic criteria, thus broadening the reach of preventive strategies (13).

The construction of PRS relies on genome-wide association studies (GWAS), which scan the genomes of large populations to identify single nucleotide polymorphisms (SNPs) associated with disease (8). However, most GWAS to date have been conducted in populations of European ancestry, meaning that PRS derived from these studies may not be well-calibrated for individuals from other backgrounds, such as Māori or Pacific peoples. This can perpetuate health inequities unless PRS are recalibrated for the target population.

Contemporary PRS are now more powerful than many single traditional risk factors and, in some cases, are as accurate as established cumulative risk scores. Importantly, a single gene chip can generate multiple PRS for a wide range of complex conditions, making genomic screening highly cost-effective and valuable across the spectrum of health care (14,15).

### 1.4 Network Medicine and Epistasis

Traditional PRS models use a linear additive model for estimating risk, which sums the total number of SNPs associated with disease, each multiplied by their attributed odds ratio. This method ignores the complex interactions between genes and proteins.

Network medicine and graph-based computational methods represent a new frontier in understanding complex disease. By modelling genes, proteins, and pathways as interconnected networks, these approaches can capture the interplay between genetic and environmental factors, identify disease modules, and predict the functional impact of genetic variants (16–18).

The NeEDL (Network Medicine-based Epistasis Detection in Complex Diseases) framework uses quantum computing-informed algorithms to identify higher-order SNP-SNP interactions that contribute to disease risk beyond simple additive effects (19).

Knowledge graphs and explainable geometric learning further enhance the interpretability and clinical utility of genetic risk models (20). The “Holo-Q” nomenclature reflects both the information-theoretic, holographic framing of omnigenic architecture (risk distributed across interconnected networks) and the quantum-informed discovery of epistatic modules.

### 1.5 Rationale for a Custom Array

Despite advances in genomic technology, several gaps remain:

1. Underdiagnosis and undertreatment of FH and high polygenic risk, especially in Māori, Pacific, and other non-European populations.
2. Limited integration of genetic testing into routine clinical care, due to cost, complexity, and lack of tailored resources.
3. Insufficient use of advanced computational methods to interpret genetic data and guide personalised treatment.
4. Persistent health inequities in cardiovascular outcomes, driven by social, economic, and biological factors.

International centres, such as the National Health Service (NHS), have already enrolled 2.1 million patients for PRS in the Our Future Health study, demonstrating the urgent need for New Zealand to implement clinical genomics (21). A single integrated test that accounts for geographic and ancestral diversity would mitigate complexity while providing a screening tool for CVD detection and personalised management.

This study describes the design and validation of the Holo-Q Omniscan Waka Te Ira, a custom Illumina Global Screening Array (GSA) v3 enriched with variants relevant to the New Zealand population. This array integrates monogenic FH detection, CAD polygenic risk assessment, and network medicine-derived epistasis content in a single research-use-only (RUO) assay.

## 2. Methods

### 2.1 Base Platform

The Illumina Global Screening Array-24 v3.0 (GSA v3) was selected as the base platform. The GSA v3 contains approximately 654,000 markers and allows for the addition of up to 50,000 custom bead types (∼43,000 custom SNPs). The platform offers >99% call rate and >99.9% reproducibility, optimised for high throughput on the iScan System (∼5,760 samples/week; ∼1.3 minutes per sample scan time) (22). The legacy GSA v3 was selected over newer v4/EX workflows to optimize clinical content and cost. Crucially, the v3 backbone contains significantly more baseline ClinVar variants (∼45,998) than the v4 keeps population screening below a $100 threshold. Although EX technology offers faster laboratory turnaround and lower DNA input, the v3 platform maintains highly scalable robotic throughput (5,700 samples/week), justifying the trade-off of processing speed for richer clinical utility and affordability.

### 2.2 Custom Content Design

The custom content layer (∼50,000 bead types) was designed to augment the GSA v3 backbone with cardiovascular-focused depth. Content was selected from multiple sources:

#### 2.2.1 FH Variants (LDLR, APOB, PCSK9)

For the three primary FH genes, variants were systematically compiled from multiple complementary sources to maximise completeness and clinical relevance. These included the Canterbury Health Laboratories LDLR variant list (July 2024), comprising 246 LDLR mutations; peer-reviewed publications reporting FH-associated variants in New Zealand and related populations (6,23–26); the ClinVar database, from which 4,085 LDLR, 1,277 PCSK9, and 3,643 APOB mutations were extracted; and the LOVD database, contributing an additional 2,414 LDLR, 355 PCSK9, and 1,261 APOB mutations. This multi-source aggregation strategy ensured comprehensive representation of both locally reported and internationally curated pathogenic variants across the three principal FH genes.

In total, 13,035 mutations were obtained for all three genes from all sources. Of these, only 7,065 are unique SNPs with known genome coordinates and known alternative and reference alleles. Of these, 348 overlapped with SNPs for the populations described earlier. Consequently, 6,717 SNPs from databases were incorporated into the array design.

#### 2.2.2 Additional Cardiovascular and Pharmacogenomic Genes

The array was further enriched with clinically relevant cardiovascular and pharmacogenomic content. Specifically, we incorporated variants from 68 genes included in the MyCode Results Reported panel, 12 key pharmacogenes representing established ADME/CPIC loci, and seven additional genes implicated in familial hypercholesterolemia or related lipid disorders, namely LDLRAP1, ABCG5, APOE, LIPA, ABCA1, LPL, and ANGPTL3. This expanded gene set was selected to provide broader coverage of cardio-metabolic pathways, rare dyslipidaemias, and pharmacogenetic determinants relevant to lipid-lowering therapy and cardiovascular risk management.

For these 87 genes, all pathogenic and likely pathogenic SNPs were extracted from ClinVar and dbSNP databases, yielding 29,947 SNPs. After filtering for design compatibility, 14,005 SNPs were included.

#### 2.2.3 Copy Number Variation (CNV) Support

All pathogenic and likely pathogenic CNVs for the 87 genes were downloaded from the dbVAR database, yielding 5,480 CNVs. For CNV detection using PennCNV (which requires at least 3 SNPs), additional probes were added for CNVs with insufficient coverage. A total of 5,845 SNPs were added for CNV detection, with 2,183 specifically added to ensure all CNVs >3bp were covered by at least 3 probes.

#### 2.2.4 CAD Epistasis Content (NeEDL)

A total of 5,232 SNPs were included from the NeEDL (Network Medicine-based Epistasis Detection) framework, prioritising higher-order CAD interactions supported by biological networks and replication analyses. These SNPs represent epistatic interaction modules identified using quantum computing-informed algorithms (19).

#### 2.2.5 PRS Enrichment

To support polygenic risk assessment, 14,806 rare variants were prioritised from the validated CAD PRS model PGS003725 (https://www.pgscatalog.org/score/PGS003725/). For each SNP in the model, the maximum population frequency was obtained using the Ensembl VEP tool, and the rarest SNPs were selected to fill the array to the maximum capacity of 50,000 beads. Additionally, broad tagging content was included to anchor GPSMult (multi-ancestry, ∼1.2M-variant) PRS calculation by imputation.

#### 2.2.6 Māori Stakeholder Engagement and Data Sovereignty and Population-Specific Founder Variants

To improve sensitivity across diverse populations, we incorporated 289 SNPs specific to New Zealand, European, Asian, and African populations, as well as 118 variants specific to populations from Japan, Korea, Thailand, and Russia. These loci were selected based on published literature describing familial hypercholesterolaemia mutations across various ancestry groups, with the aim of enhancing detection accuracy in ancestrally diverse populations that are inadequately captured by standard reference datasets.

##### Ethics, consultation and stakeholder engagement

This approach ensures that the research is conducted in accordance with principles of Māori data sovereignty and cultural protocols, recognising genomic data as taonga and respecting the rights and interests of Māori communities in research involving indigenous populations and resources. Safeguarding intellectual and cultural property requires data sovereignty policies that align with Te Tiriti o Waitangi, Wai 262 considerations, and international conventions such as the United Nations Convention on Biological Diversity and the Nagoya Protocol. Upholding tikanga, respecting Māori aspirations, and fostering equitable partnerships ensures research benefits flow back to the communities from which they arise.

The planned validation in Māori and Pacific cohorts is a strength of the programme, provided governance and consent conditions are respected and results are reported in an equity-forward manner. Where PRS performance and clinical utility differ by ancestry, socioeconomic context, or health-system access, these factors will be transparently communicated to clinicians and patients.

Because polygenic risk score methods can perform unevenly across populations, the programme explicitly commits to ancestry-stratified performance reporting for Māori, Pacific, and other key groups where sample size permits; calibration approaches appropriate to each group; transparent communication of limitations where sample sizes constrain precision; and avoidance of deployment approaches that could amplify existing health inequities.

##### Equity and avoidance of harm

Discussions with government regarding the programme have emphasised the importance of preserving data sovereignty for nations and communities using the chip, and have included consideration of benefit-sharing frameworks and iwi leadership in related initiatives such as the New Zealand Digital Pharmacopoeia.

The research programme is committed to ensuring that benefits flow back to the communities from which data and knowledge arise. This includes Māori and Pacific co-authorship and leadership roles where these populations materially contribute to validation and implementation; capacity building through training pathways and technical roles; Māori-serving clinical pathways with evaluation endpoints chosen in consultation with Māori clinical leadership; and commitments regarding affordability and access in deployment settings relevant to Māori communities.

##### Benefit sharing and reciprocity

Key governance principles include: Māori oversight of decisions regarding secondary uses of Māori genomic data; tiered access models with appropriate controls; defined processes for data retention and participant withdrawal; and clarity regarding governance of derived artefacts such as imputed genotypes, polygenic risk scores, and computational models.

Collected human genomic data will be stored within established institutional frameworks (including the Auckland Regional Tissue Bank and NeSI infrastructure) with appropriate governance structures in place. The programme is designed to deposit genomic data into the Rakeiora Precision Medicine Research Platform, a Genomics Aotearoa pathfinder project that has developed a national-scale computational infrastructure with indigenous data sovereignty attributes at its core (37). Rakeiora uses a ‘walled garden’ approach where all genomic and health data analysis occurs within a secure computational environment with rigorous permission systems, precise tracking of data access, and appropriate permissioning before results can be exported.

Genomic data are closely connected to whakapapa (genealogy) and identity, and therefore raise distinct obligations for governance, culturally safe engagement, and the protection of Māori rights and interests. Data, information, or knowledge that is about, from, or connected to Māori is considered taonga, and accordingly principles of Māori data sovereignty and governance inform the storage and use of data generated through this programme.

##### Māori data sovereignty and governance

The Holo-Q platform was given a Māori name—Waka Te Ira—reflecting its positioning within an Aotearoa New Zealand context and signaling commitment to Māori partnership. The chip’s performance and associated polygenic risk score models are being benchmarked using cohorts that include Māori and Pacific participants who have consented to genotyping, with ancestry-stratified performance reporting to ensure transparency about model validity across populations.

These consultations established several key principles: Māori investigators and advisors would be engaged throughout the programme; knowledge generated from the research would be applied to benefit Māori communities; the platform would be designed to serve the genetic architecture of New Zealand’s diverse population, including Māori and Pacific peoples; and validation would include Māori and Pacific cohorts to ensure equitable analytic performance rather than extrapolation from non-representative populations.

Meaningful engagement with Māori represents an ongoing process that ideally establishes enduring, trusted relationships and continues throughout the research lifecycle. In the development and validation of the Holo-Q Waka Te Ira Omniscan genotyping array, the research team drew upon established relationships with Māori academics, and scientists who were consulted regarding study design, implementation, and governance considerations.

### 2.3 Final Array Composition

The final custom design file contained 43,437 SNPs utilising 50,000 beads. Combined with the GSA v3 backbone, the total array content provides comprehensive genome-wide coverage with focused cardiovascular depth. In total, 47,027 SNPs were included on the array, of which 3,590 were already present in the GSA v3 backbone, meaning that 43,437 SNPs were newly added as part of the custom content.

### 2.4 Validation Datasets

Validation was performed using data from the UK Biobank (UKBB):

1. Imputed Genotype Data (TOPMed): Obtained from the UK Biobank DNAnexus. The dataset comprised 487,180 samples in BGEN format.
2. DRAGEN WGS Dataset: Joint-called Whole Genome Sequencing data from 487,180 samples.
3. GraphTyper WGS Dataset: Population-level genome variants from 200,842 samples.

SNP frequency analysis was performed using qctool v. 2.2.0. On the DNAnexus server, qctool software was compiled into a qctool applet and executed using the command ‘dx run’.

#### 2.4.1 High-Performance Compute (HPC) Cybersecurity Measures

For computational efficiency a hybrid system was used i.e. DNA Nexus (chromosomes 4-9) and Cerebro server (chromosomes 1-3, 10-22), a custom-built workstation equipped with a 24-core AMD Ryzen Threadripper 3960X processor, 128 GB of DDR4 RAM, and an NVIDIA GeForce RTX 3060 GPU (12 GB VRAM). Local data storage was provided by an 18 TB hard drive. Given the sensitive nature of the UK Biobank data utilised for the Holo-Q Omniscan validation, stringent cybersecurity measures were implemented on the Cerebro HPC (custom-built workstation at Te Whatu Ora). Data-at-rest security was achieved through full-disk encryption, which was immediately deployed on the primary 18 TB data drive. The encryption keys are securely managed within the Trusted Platform Module (TPM) to facilitate automated system restarts. For remote network access, a more robust solution was established using Cloudflare Zero Trust, disallowing direct SSH and routing all remote connections through a secure, firewalled intermediary. Access for all users was mediated by a mandatory two-factor authentication (2FA) process. These information security controls are part of the platform’s commitment to compliance, aimed at meeting the information security policy requirements, such as the ISO 27001 standard, mandated by the UK Biobank Material Transfer Agreement (MTA).

### 2.5 PRS Model Benchmarking

To evaluate the performance of polygenic risk scores achievable with the Holo-Q array, we benchmarked multiple PRS models using simulated Holo-Q data.

#### 2.5.1 Simulated Holo-Q Data Generation

We extracted Holo-Q loci from the WGS dataset. For genotype imputation, to reduce computational burden, we selected a subset of 6,000 samples and augmented this set with additional individuals to ensure that all variants were polymorphic. Pre-phasing was performed using Eagle, followed by imputation with Minimac4, using the 1000 Genomes Project Phase 3 (30× coverage) panel as the reference. The imputed dataset comprised approximately 50 million variants genome-wide.

#### 2.5.2 CAD PRS Models

From the PGS Catalog (https://www.pgscatalog.org/trait/EFO_0001645/), 78 PGS models for coronary artery disease were identified. After excluding 16 models where only effect alleles were provided (no alternative alleles), models were further selected based on two criteria: development using more than 1 million SNPs and training in populations beyond solely European ancestry, resulting 10 models were evaluated. CAD case status was defined by ICD-10 codes (I21, I22, I23, I24.1, I25.2) and OPCS-4 codes (K40.1-40.4, K41.1-41.4, K45.1-5, K49.1-49.2, K49.8-49.9, K50, K75.1-75.4, K75.8-75.9), identifying 34,082 cases (6.97% of the cohort).

#### 2.5.3 LDL-C PRS Models

For LDL cholesterol, 121 models were identified from the PGS Catalog (https://www.pgscatalog.org/trait/EFO_0004611/). After excluding 5 models without reference alleles, the same selection criteria (more than 1 million SNPs and multi-ancestry training) were applied, yielding 10 models were evaluated. LDL values were extracted from UKBB field p30780_i0, yielding 406,213 samples with valid measurements (mean = 3.54 mmol/L, SD = 0.89).

#### 2.5.4 Statistical Analysis

For CAD, the area under the receiver operating characteristic curve (AUC-ROC) was calculated comparing predicted PGS values against true CAD case/control status. Models were evaluated both unadjusted and adjusted for covariates (sex, age, first four genetic principal components, and genotyping batch). For LDL-C, the coefficient of determination (R²) and Pearson correlation coefficient were calculated.

### 2.6 FH Variant Validation Against ClinVar

The ClinVar database contained 7,404 mutations associated with familial hypercholesterolemia. Of these, 1,410 SNPs were classified as pathogenic or likely pathogenic, occupying 1,112 unique genomic positions. The array design was compared against this reference to assess coverage of clinically actionable FH variants.

## 3. Results

### 3.1 SNP Recovery in UK Biobank Imputed Data

Out of the 43,437 unique SNPs included in the custom design, 22,483 (51.8%) were identified in the UKBB imputed data. The mean minor allele frequency (MAF) for identified SNPs was 0.188, with a median of 0.163. Among these, 6,741 SNPs had MAF < 0.05, and 5,441 SNPs had MAF < 0.01.

The results demonstrate that the vast majority of SNPs specific to New Zealand and Asian populations are absent in UKBB, likely due to the underrepresentation of these populations in the dataset. This finding validates the rationale for including population-specific content in the custom design. In contrast, most SNPs from the CAD SNPs and PRS SNPs categories were identified in UKBB with high MAF values, as expected given that UKBB data were used in constructing these prediction models.

### 3.2 Participant-Level Variant Distribution

Analysis of SNP distribution among UKBB participants revealed that each participant carried an average of 9,287 SNPs (out of 43,437 included in the design and 22,483 detected in UKBB) in either heterozygous or homozygous state. The minimum number of variants per participant was 8,269, while the maximum was 10,583. On average, participants carried 5,688 SNPs in heterozygous state and 3,599 SNPs in homozygous state.

### 3.3 Validation of FH Pathogenic Variants

The array design successfully incorporated 938 (84%) of the 1,112 unique pathogenic/likely pathogenic FH SNPs listed in ClinVar. The standard GSA v3 array contains 742 of these variants; thus, our custom design added 196 unique pathogenic variants, representing a 26.4% expansion in FH coverage.

The remaining 174 pathogenic/likely pathogenic FH-associated SNPs not included in the design may reflect: (1) SNPs added to ClinVar after the database was extracted for array design (August 2024), or (2) differences between the manifest file for the GSA v3 array downloaded from the Illumina website and the array version uploaded in Illumina DesignStudio.

### 3.4 WGS Validation and Rare Variant Detection

The 938 FH variants were screened against UK Biobank whole-genome sequencing (WGS) datasets generated using both the GraphTyper and DRAGEN pipelines. In the GraphTyper WGS dataset (n = 200,842), 176 of the 742 FH-associated SNPs present on the standard GSA v3 array were detected, with a mean minor allele frequency (MAF) of 0.002 (SD = 0.028; range 0.00002–0.371). Of these, 175 variants (99.4%) exhibited MAF < 0.01, and 172,723 individuals (86%) carried at least one mutant allele. Among the 196 newly added custom FH SNPs, 53 were detected in the GraphTyper dataset, with a mean MAF of 0.000014 (SD = 0.016; range 0.00002–0.000135), and only 290 participants (0.14%) harboured mutations in these loci.

In contrast, analysis of the DRAGEN joint-called WGS dataset (n = 487,180) identified only a single variant (chr1:161223893G>A in the LDLR gene). This unexpectedly low detection rate likely reflects technical factors rather than true absence of variants, as the DRAGEN pipeline applies stringent quality filters that may systematically exclude ultra-rare variants (MAF < 0.00001). In addition, potential coordinate discrepancies between the DRAGEN reference build and the positional annotations used in the array design may have contributed to variant misalignment and under-detection.

Excluding the high-frequency APOA2 variant (rs5082, alternative allele frequency 0.63), pathogenic or likely pathogenic FH-associated SNPs were observed in only 4,678 of 200,000 UKBB participants (2.34%) using standard GSA v3 content. The custom design identified an additional 290 carriers (0.14%)—high-risk individuals who would have been missed by the standard array.

### 3.5 CAD PRS Model Performance

Benchmarking of 10 CAD PRS models using simulated Holo-Q imputed data revealed substantial variation in predictive accuracy. The inclusion of covariates (sex, age, PC1-4, genotyping batch) significantly improved model performance, increasing the mean AUC-ROC from 0.626 (unadjusted) to 0.764 (adjusted).

The highest-performing models (PGS004697, PGS004696, PGS003356) were trained on datasets that included UKBB, potentially inflating their apparent accuracy due to overfitting. Among models not trained on UKBB data, PGS003725 achieved the highest accuracy (AUC 0.786 adjusted), making it the recommended model for unbiased clinical implementation. This model was therefore selected for rare variant enrichment in the array design.

### 3.6 LDL-C PRS Model Performance

Benchmarking of 10 LDL-C PRS models revealed that covariate adjustment had minimal effect on model performance (mean R² increase from 0.092 to 0.093). Model PGS002337 consistently demonstrated the highest predictive accuracy (r = 0.190 adjusted), followed by PGS003037 (r = 0.178 adjusted).

Notably, PGS000888—reported in the literature as the top-performing LDL model (R² = 0.176)—yielded substantially lower accuracy in our validation (r = 0.099). This discrepancy likely reflects differences in population structure: the published performance was achieved in a European-only sample, whereas our analysis included individuals of diverse ancestries within UKBB. This observation aligns with known challenges in trans-ancestry polygenic prediction.

### 3.7 Imputation Performance

Simulation of Holo-Q content imputed against the 1000 Genomes Phase 3 (30x) reference panel demonstrated robust imputation performance. Across all chromosomes, the imputed data yielded approximately 53 million variants, with high concordance between imputed and WGS-derived genotypes for common and low-frequency variants (MAF ≥1%). The summary of imputed variant counts by chromosome is presented in Table 7.

**Table 1.** Summary of Custom SNP Content by Category.

| Category | SNPs in Design | Purpose |
| --- | --- | --- |
| LDLR, APOB, PCSK9 | 6,717 | Comprehensive FH gene coverage |
| Additional Genes (MyCode + FH-related + pharmacogenes) | 14,005 | Broader cardio-metabolic coverage |
| CNV Support | 5,845 | Array-based CNV detection |
| CAD Epistasis (NeEDL) | 5,232 | Higher-order interaction content |
| PRS Enrichment (PGS003725) | 14,806 | Rare PRS variant coverage |
| NZ, European, Asian, African | 289 | Population-specific variants |
| Japan, Korea, Thailand, Russia | 118 | East Asian founder variants |

**Table 2.** Recovery of Custom SNP Categories in UK Biobank.

| Category | In Design | Found in UKBB | Mean MAF | Median MAF |
| --- | --- | --- | --- | --- |
| NZ-specific | 289 | 44 (15.2%) | 0.028 | 0.00002 |
| Asian-specific | 130 | 10 (7.7%) | 0.00002 | 0.00002 |
| LDLR, PCSK9,<br>APOB SNPs | 6,717 | 910 (13.5%) | 0.012 | 0.00002 |
| Additional<br>Genes | 14,005 | 1,550 (11.1%) | 0.082 | 0.048 |
| For CNV | 5,845 | 443 (7.6%) | 0.005 | 0.00002 |
| CAD SNPs | 5,232 | 4,704 (89.9%) | 0.261 | 0.260 |
| PRS SNPs | 14,806 | 14,802 (99.9%) | 0.194 | 0.170 |

**Table 3.**
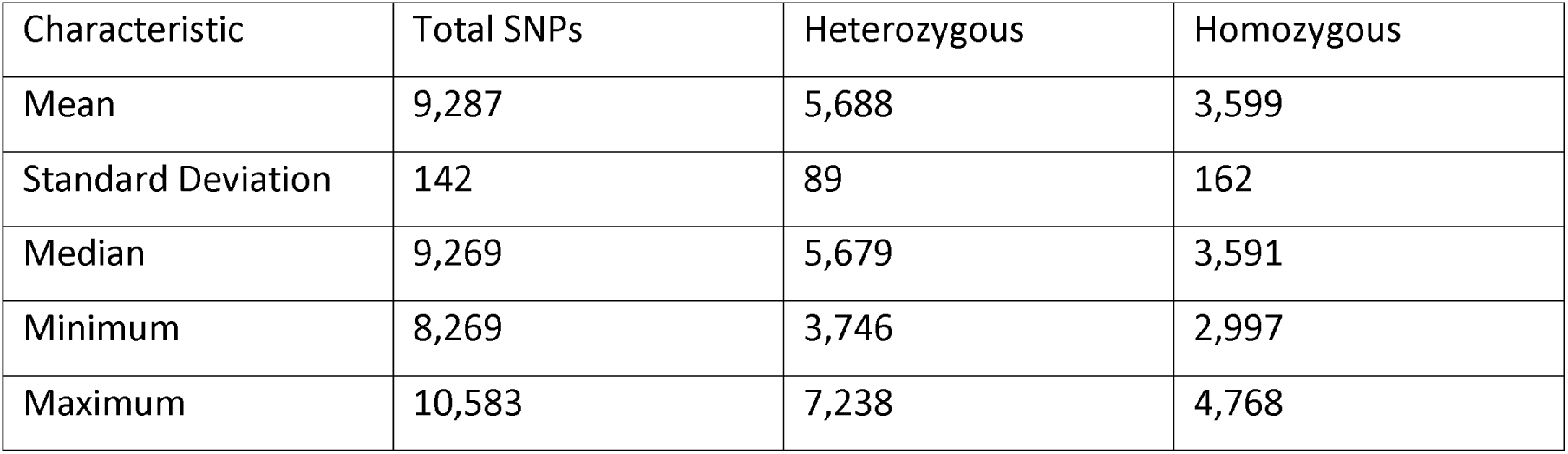
Distribution of SNP Counts per UK Biobank Participant.

**Table 4.** Detection of FH Variants Across WGS Datasets.

| Content | Total SNPs | Found in<br>DRAGEN | Found in<br>GraphTyper | Patients with<br>SNP |
| --- | --- | --- | --- | --- |
| Custom Design<br>(New) | 196 | 0 | 53 | 290 (0.14%) |
| Standard GSA v3 | 742 | 1 | 176 | 4,678 (2.34%) |

**Table 5.** Top-Performing CAD PRS Models (Holo-Q Simulation).

| PGS ID | Method | AUC (Raw) | AUC (Adjusted) | AUC (Adj+PCA) |
| --- | --- | --- | --- | --- |
| PGS004697 | PRS-CSx | 0.731 | 0.811 | 0.825 |
| PGS004197 | PRS-CSx | 0.742 | 0.819 | 0.823 |
| PGS000329 | LDpred | 0.730 | 0.815 | 0.819 |
| PGS004696 | PRS-CSx | 0.718 | 0.802 | 0.817 |
| PGS003356 | LDpred | 0.725 | 0.814 | 0.817 |
| PGS004200 | PRS-CSx | 0.723 | 0.809 | 0.813 |
| PGS005091 | PRS-CSx | 0.710 | 0.800 | 0.812 |
| PGS003725* | LDpred2 | 0.665 | 0.784 | 0.786 |
| PGS000013 | LDpred | 0.627 | 0.763 | 0.765 |
| PGS000018 | metaPRS | 0.619 | 0.763 | 0.767 |

\*PGS003725 was selected for array enrichment as it was not trained on UKBB data, providing unbiased performance estimates.

**Table 6.** Top-Performing LDL-C PRS Models (Holo-Q Simulation).

| PGS ID | Method | r (Raw) | r (Adjusted) | r (Adj+PCA) |
| --- | --- | --- | --- | --- |
| PGS002337 | BOLT-LMM | 0.175 | 0.185 | 0.190 |
| PGS003037 | DBSLMM | 0.164 | 0.175 | 0.178 |
| PGS002703 | SBayesR | 0.153 | 0.162 | 0.165 |
| PGS001933 | bigstatsr | 0.149 | 0.158 | 0.160 |
| PGS003032 | DBSLMM | 0.144 | 0.155 | 0.158 |
| PGS003038 | PRS-CS(auto) | 0.138 | 0.146 | 0.154 |
| PGS003033 | PRS-CS(auto) | 0.138 | 0.147 | 0.153 |
| PGS003029 | Lassosum | 0.133 | 0.144 | 0.147 |
| PGS000888 | PRS-CS | 0.084 | 0.093 | 0.099 |
| PGS003785 | LDpred2 | 0.051 | 0.061 | 0.067 |

**Table 7.** Summary of Imputed Variants by Chromosome.

| Chromosome | HoloQ content<br>(DRAGEN-WGS<br>matched) | Imputation total<br>(DRAGEN-WGS<br>matched) |
| --- | --- | --- |
| 1 | 49,083 | 4,028,743 |
| 2 | 53,928 | 4,357,823 |
| 3 | 41,793 | 3,659,541 |
| 4 | 39,988 | 3,551,569 |
| 5 | 38,019 | 3,353,704 |
| 6 | 42,885 | 3,157,937 |
| 7 | 34,532 | 3,015,503 |
| 8 | 32,995 | 2,852,530 |
| 9 | 26,773 | 2,222,461 |
| 10 | 30,496 | 2,541,124 |
| 11 | 30,466 | 2,565,620 |
| 12 | 29,371 | 2,452,680 |
| 13 | 21,762 | 1,744,681 |
| 14 | 21,076 | 1,623,286 |
| 15 | 18,967 | 1,491,316 |
| 16 | 20,448 | 1,645,030 |
| 17 | 18,874 | 1,466,443 |
| 18 | 17,462 | 1,443,913 |
| 19 | 16,085 | 1,157,777 |
| 20 | 14,518 | 1,141,310 |
| 21 | 8,445 | 703,490 |
| 22 | 8,983 | 701,875 |
| X | 24,483 | 1,668,480 |
| Y | 3,211 | 3,211 |
| MT | 970 | 970 |

## 4. Discussion

This study presents the design and validation of the Holo-Q Omniscan Waka Te Ira, a custom genotyping array purpose-built for integrated cardiovascular genomics in New Zealand’s multi-ethnic population. The array represents a significant advance over standard genotyping platforms by combining monogenic FH detection, state-of-the-art polygenic risk assessment, and network medicine-derived epistasis content in a single assay.

### 4.1 Enhanced FH Detection

By adding 196 pathogenic variants not present on the standard GSA v3 array, we achieved a 26.4% expansion in coverage of clinically actionable FH variants. In the UKBB proxy analysis, this translated to identifying 290 additional carriers of rare, high-impact mutations—individuals who would have been missed by standard arrays. Given that early detection and treatment of FH can reduce MACE risk by up to 80% (3), this enhanced detection capability has significant clinical implications.

A published study using a customised Illumina GSA FH array (636 variants) demonstrated overall sensitivity of 94.7% (98.2% when only variants present on the array were considered) and CNV sensitivity of 89.4% (5). This supports the feasibility and clinical utility of FH-oriented array designs on the Infinium platform.

### 4.2 Polygenic Risk Assessment

The near-perfect recovery of PRS and CAD SNPs (99.9% and 89.9%, respectively) ensures that the array can effectively generate polygenic risk scores. Our benchmarking identified PGS003725 as the optimal model for unbiased clinical implementation, achieving an AUC of 0.786 when adjusted for covariates. This model was specifically selected because it was not trained on UKBB data, avoiding the overfitting that inflated the apparent performance of other models.

The integration of GPSMult-tagging content enables multi-ancestry PRS calculation through imputation. A 10-year serial CCTA study demonstrated that higher CAD PRS (GPSMult) is associated with greater atheroma progression and markedly higher odds of high-risk plaque features, with PRS improving prediction of non-calcified plaque progression beyond conventional risk factors (27). This clinical evidence supports the value of PRS enrichment in the array design.

### 4.3 Network Medicine and Epistasis

The inclusion of 5,232 NeEDL-identified epistasis SNPs represents a novel approach to capturing higher-order genetic interactions. Traditional PRS models assume additive effects, ignoring the complex interplay between genes and pathways that characterises the omnigenic architecture of complex diseases (16–21). By embedding network medicine-derived content, the Holo-Q array enables next-generation risk models that better reflect the interconnected basis of cardiovascular disease.

The “holographic” framing reflects the information-theoretic concept that risk is distributed across interconnected networks—the whole is encoded in every part. This design philosophy moves beyond simple variant counting toward a more biologically informed approach to genetic risk assessment.

### 4.4 Addressing Health Inequities

The poor recovery of NZ-specific (15.2%) and Asian-specific (7.7%) variants in the UK Biobank serves as a “negative control,” demonstrating that reliance on standard, Eurocentric datasets fails to capture the genetic architecture of New Zealand’s diverse population. This finding strongly supports the deployment of this custom array for local implementation.

The inclusion of 289 variants specific to New Zealand, European, Asian, and African populations, along with 118 variants specific to Japan, Korea, Thailand, and Russia, addresses a critical gap in current genomic tools. These population-specific variants are essential for equitable genomic medicine, ensuring that diverse populations receive high-quality genetic risk assessment.

### 4.5 Comparison with Other Arrays

The Holo-Q Omniscan differs from other available arrays in several key respects: Versus GDA Enhanced (co-developed with Allelica): GDA Enhanced is optimised for broad, multi-ancestry PRS deployment across many diseases. Holo-Q emphasises cardiovascular depth with extensive FH pathogenic content (including CNVs), targeted population-specific founder variants, NeEDL epistasis SNPs, and cardio-metabolic/PGx loci while still supporting multi-ancestry CAD PRS.

Versus Excalibur (Our Future Health): The Excalibur array, designed for the UK’s Our Future Health (OFH) program, represents the current state-of-the-art for biobank-scale clinical screening. A major design priority for Excalibur was improving trans-ancestry risk prediction by adding hundreds of thousands of bespoke variants optimized for global diversity, driven primarily by population allele frequencies and standard linear GWAS methods.

While Excalibur is a broad, discovery-oriented, pan-disease tool containing over 692,000 variants, Waka Te Ira utilizes a fundamentally distinct, targeted design philosophy. A comparative analysis reveals that while the arrays share a massive foundational footprint—overlapping by 525,613 variants from the standard GSA v3 backbone (covering roughly 80% of the GSA baseline)—Holo-Q diverges significantly in its custom architecture.

Of the 43,437 custom variants added to Holo-Q, only 4,643 overlap with the Excalibur array. The remaining 38,152 custom variants are entirely unique to Holo-Q. This distinct profile is defined by the intentional inclusion of regionally validated FH variants (sourced directly from Canterbury Health Laboratories) and NeEDL SNPs involved in genetic epistasis. Rather than relying solely on global allele frequencies, this unique custom content is anchored in human biology—targeting specific interacting macromolecular networks to capture higher-order disease relationships that linear, additive models typically omit.

Ultimately, while the 38,152 unique custom variants optimize Holo-Q for network-informed cardiovascular risk and geographically tailored detection, the extensive backbone overlap (>525,000 SNPs) ensures structural complementarity with massive population efforts like Excalibur. This shared genomic footprint inherently opens the door to future international collaborations and multi-cohort analyses via secure, federated methods.

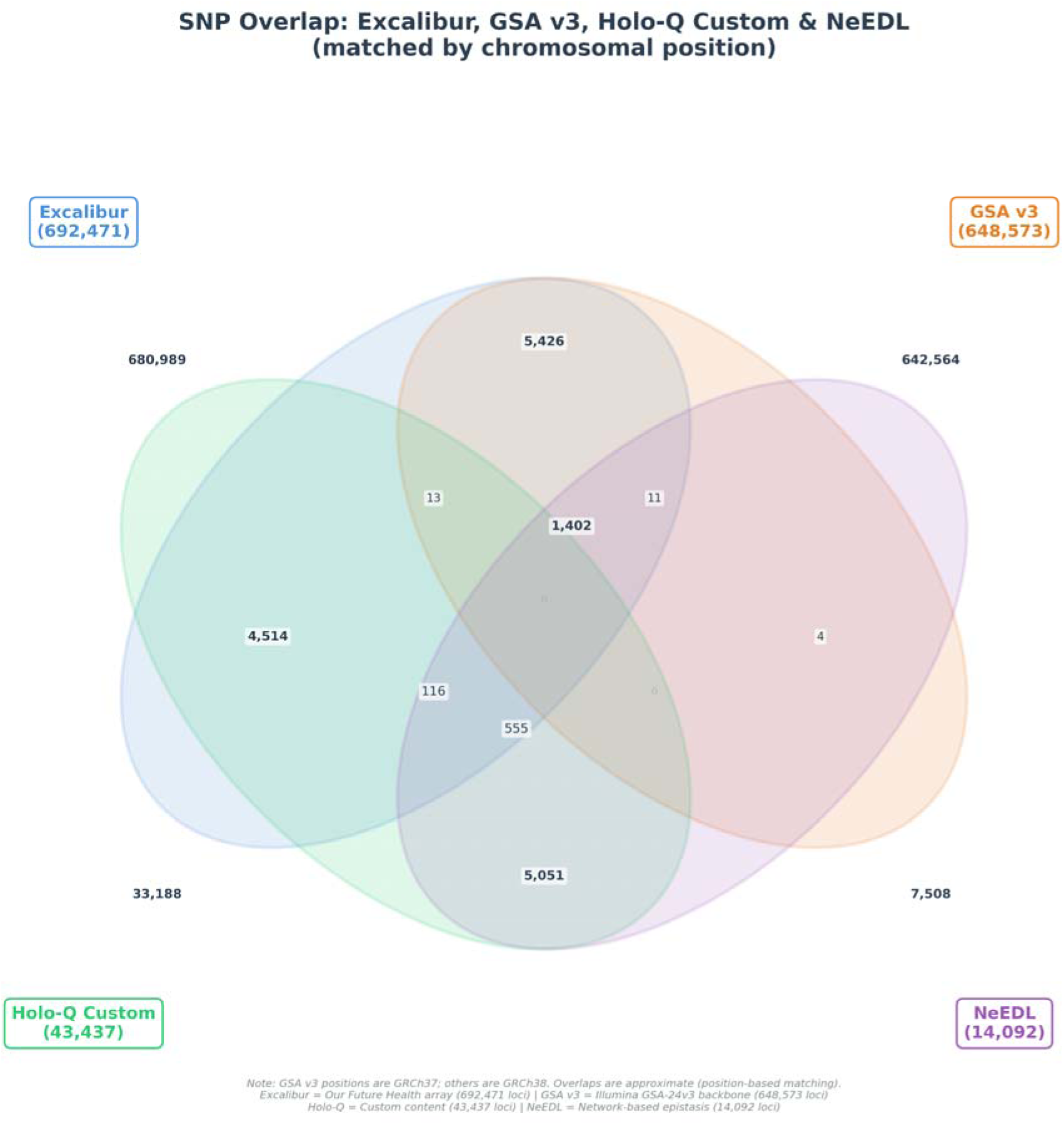

### 4.6 Limitations

Several limitations should be acknowledged. First, validation was performed using UK Biobank data, which underrepresents the populations of greatest interest (Māori, Pacific, Asian). Local validation in New Zealand cohorts is essential before clinical implementation. Second, the DRAGEN WGS analysis yielded unexpectedly low variant detection, likely due to technical factors rather than true absence of variants. Third, 174 pathogenic/likely pathogenic FH variants from ClinVar were not included in the design, potentially due to database updates after the design freeze or manifest discrepancies. Finally, the epistasis content from NeEDL requires further validation in independent cohorts to confirm its clinical utility.

While the Holo-Q array demonstrates excellent performance for common variants and pre-specified pathogenic mutations, inherent limitations of array-based genotyping must be acknowledged. Arrays cannot detect variants not included in the design—a critical limitation for ultra-rare or population-private variants that may be clinically significant but absent from reference databases (28). Furthermore, rare variants on arrays may generate more false positives than true positives due to probe interference at adjacent positions (29). Ultra low-coverage WGS (0.4x) has demonstrated superior detection of rare variants (MAF <1%) compared to GSA arrays, with higher non-reference concordance (0.90 vs 0.88) (30).

These limitations support a two-tier diagnostic strategy: (1) initial array-based screening for PRS calculation and detection of known pathogenic FH variants, followed by (2) targeted next-generation sequencing for individuals with either a positive mutation finding requiring confirmation, or those with high monogenic probability (low LDL-C PRS but severely elevated LDL-C levels). This approach maximizes cost-effectiveness while ensuring that individuals with potential novel or population-specific pathogenic variants are not missed by array-based screening alone.

### 4.7 Future Directions

This array is designed to support a three-year prospective study targeting ACS patients admitted to coronary care units in New Zealand. The study will:

1. Determine the prevalence and detection accuracy of monogenic and polygenic FH in a contemporary ACS cohort
2. Assess the incremental predictive value of advanced PRS methodologies, including knowledge graphs and network science
3. Evaluate the impact of genomics-guided care on clinical outcomes (LDL-C reduction, MACE), prescribing behaviour, medication adherence, and patient satisfaction
4. Examine the cost-effectiveness and health equity implications of implementing genomics-guided screening

Additionally, the study will bank whole-blood PAXgene RNA tubes and store DNA for methylation arrays, creating a multi-omics resource (genotype, transcriptome, epigenome) that can be analysed within the same knowledge-graph framework.

Layering gene-expression and methylation information onto the network enables modelling of higher-order epistatic interactions, further refining polygenic risk.

## 5. Conclusion

The Holo-Q Omniscan Waka Te Ira represents a purpose-built solution for integrated cardiovascular genomics in New Zealand. By combining comprehensive FH variant detection (26.4% expansion over standard arrays), robust polygenic risk assessment (AUC 0.786 for CAD), and novel network medicine-derived epistasis content, the array bridges the gap between standard genotyping and the specific needs of a multi-ethnic population.

The validation against UK Biobank data confirms excellent performance for common variants while demonstrating that population-specific content is essential for equitable genomic medicine. This tool is poised to facilitate precision cardiovascular care, enabling earlier detection of high-risk individuals, personalised treatment strategies, and ultimately, reduced cardiovascular morbidity and mortality in New Zealand.

## Data Availability

All data produced in the present study are available upon reasonable request to the authors

## Acknowledgments

This research has been conducted using the UK Biobank Resource under Application Number [93783]. We thank the participants and researchers who contributed to UK Biobank. We gratefully acknowledge Drs. Weerapan Khovidhunkit, Kanya Suphapeetiporn, and Poranee Ganokroj of King Chulalongkorn Memorial Hospital for sharing the Thai exome FH variants placed on the Holo-Q chip. We affirm that this genetic data is subject to the same strict data sovereignty protections as indigenous data. Thanks to Cherry Weng for organizing the figures, tables and transferring information. We also thank Prof. Philip Wilcox, Dr. Nikki Earle, Prof Murray Cox and Prof. Rinki Murphy for their support and critical review of the manuscript. Thanks to Matt Weston at Prompt Innovation for his work in cybersecurity on Cerebro. We also thank Kevin Smith for his diligence with managing DNA within the Waitematā biobank and Andrew Laurie at Canterbury Health Laboratories for sharing FH variants, under approval of the national FH registry’s ethics committee.

