## Supplemental for "A Custom Global Screening Array for Integrated Familial Hypercholesterolemia Detection and Polygenic Risk Assessment in a Multi-Ethnic New Zealand Population"

### Supplementary Methods

**Holo-Q Omniscan Validation Study: Pipeline Architecture and Clinical Implementation**

#### S1. Bioinformatics Pipeline Overview

The Holo-Q Omniscan validation study employs a comprehensive bioinformatics pipeline for genotype processing, imputation, and polygenic risk score (PRS) calculation. The pipeline architecture is illustrated in Supplementary Figure S1.

##### S1.1 Data Ingestion and Quality Control

Genotype data from the Holo-Q Omniscan array (Illumina iDAT format) undergoes initial quality control including:

• Sample call rate filtering (>95%)
• SNP call rate filtering (>95%)
• Hardy-Weinberg equilibrium testing (p > 1×10⁻⁵)
• Sex concordance verification
• Heterozygosity rate assessment

##### S1.2 Phasing and Imputation

Phasing is performed using SHAPEIT4 with genetic map files aligned to GRCh38. Imputation utilizes either Minimac3 or Selphi against the 1000 Genomes Project Phase 3 high-coverage reference panel (3,202 samples). The reference panel is pre-converted to M3VCF format (Minimac3) or PBWT format (Selphi) to optimize computational performance.

Computational requirements: Imputation of one sample across all 22 autosomes requires approximately 2 hours when parallelized across 22 CPU cores with ~40GB RAM.

##### S1.3 Polygenic Risk Score Calculation

PRS is calculated as a weighted sum of risk alleles:

*PRS = Σᵢ βᵢ × gᵢ*

where βᵢ represents the effect weight (log odds ratio) and gᵢ represents the dosage (0, 1, or 2) of the effect allele at locus i. SNP weights are sourced from the PGS Catalog (www.pgscatalog.org) with automatic coordinate liftover from GRCh37 to GRCh38 where required.

#### S2. Holo-Q Omniscan Array Design

The Holo-Q Omniscan array is built on the Illumina GSA v3/v4 or GDA backbone with custom content optimized for cardiovascular risk assessment and familial hypercholesterolemia (FH) detection. The array design schematic is shown in Supplementary Figure S2.

##### S2.1 Core Content Categories

| Content Category | Description |
| --- | --- |
| FH Variant Panel | Population-specific FH variants from New Zealand, Thailand, Korea, and Japan; curated from ClinVar, LOVD, CPIC, and PharmGKB |
| CAD PGS Loci | Genome-wide significant and suggestive loci for coronary artery disease polygenic scoring |
| Epistasis-Enriched Variants | Variants selected based on chromatin interaction mapping, trans-eQTL networks, enhancer-promoter loops, transcription factor networks, and epigenetic modifier interactions |
| Pharmacogenomic Markers | CPIC Level A/B variants for statin response, PCSK9 inhibitor efficacy, and other cardiovascular medications |
| Ancestry Informative Markers | SNPs for accurate ancestry inference and PRS calibration across diverse populations |

##### S2.2 Epistasis-Aware Content Selection

Beyond traditional additive PRS loci, the array includes variants selected through epistasis-aware algorithms including:

• NeEDL (Network-medicine/quantum-derived epistasis detection)
• Chromatin interaction mapping (Hi-C, ChIA-PET)
• Trans-eQTL network analysis
• Enhancer-promoter loop identification
• Transcription factor binding site networks

##### S2.3 Data Sources

Array content is derived from multiple curated sources:

**• Bioinformatics databases:** ClinVar, LOVD, CPIC, PharmGKB
**• Biobanks:** UK Biobank, Geisinger MyCode, federated clinical biobanks
**• PGS Catalog:** GPSMult, PRSMix, rare variant PRS for trans-ancestry portability
**• Pangenome resources:** Ancestral recombination graphs (ARG), genotype representation graphs (GRG)

**Supplementary Figure S2. Holo-Q Omniscan Array Design Schematic**

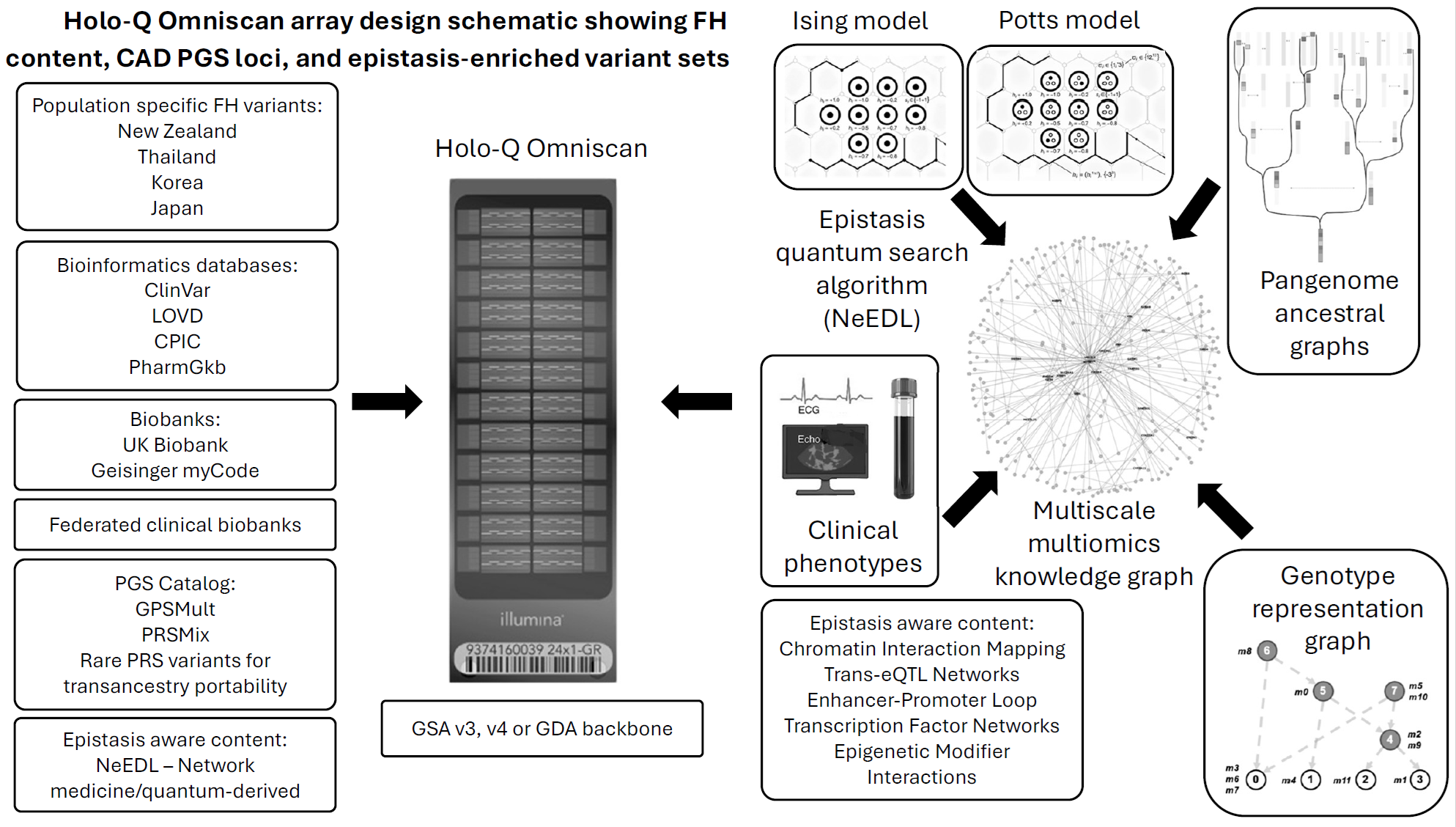

Holo-Q Omniscan array design schematic showing FH content, CAD PGS loci, and epistasis-enriched variant sets. The array integrates content from multiple data sources including population-specific FH variants, bioinformatics databases (ClinVar, LOVD, CPIC, PharmGKB), biobanks (UK Biobank, Geisinger MyCode), and the PGS Catalog. Epistasis-aware content is selected using quantum-derived algorithms (NeEDL) and network-based approaches. The design incorporates Ising and Potts model frameworks for capturing variant interactions, pangenome ancestral graphs for population diversity, and multiscale multi-omics knowledge graphs for biological context.

#### S3. Clinical Workflow and Implementation

The Holo-Q Omniscan system is designed for seamless integration into clinical workflows. The population-scale screening pipeline and clinical implementation pathway are illustrated in Supplementary Figures S3 and S4.

##### S3.1 Sample Collection and Processing

The clinical workflow comprises the following steps:

**1. Patient consent:** Electronic consent (e-consent) via tablet interface with educational video
**2. Sample collection:** Saliva or blood sample collected by trained nursing staff
**3. Genotyping:** Illumina iScan processing with custom Holo-Q Omniscan array
**4. Bioinformatics:** Automated pipeline for QC, imputation, and PRS calculation
**5. Clinical reporting:** Generation of clinician-facing and patient-facing reports
**6. Clinical review:** Results reviewed in personalized medicine clinic with multidisciplinary team

##### S3.2 Indigenous Data Governance

The pipeline incorporates robust indigenous data governance frameworks including:

• Bioculture labels for data provenance and usage rights
• Walled garden architecture for indigenous genome databases
• Encryption and blockchain-based audit trails
• Benefit-sharing mechanisms
• University research governance partnerships

##### S3.3 Ancestry and Population Stratification

Principal component analysis (PCA) is performed against 1000 Genomes superpopulations (EUR, EAS, AMR, SAS, AFR) to ensure appropriate PRS calibration. Ancestral recombination graphs (ARG) and genotype representation graphs (GRG) are used for refined ancestry inference and population-specific risk adjustment.

**Supplementary Figure S3. Population-Scale Screening Pipeline**

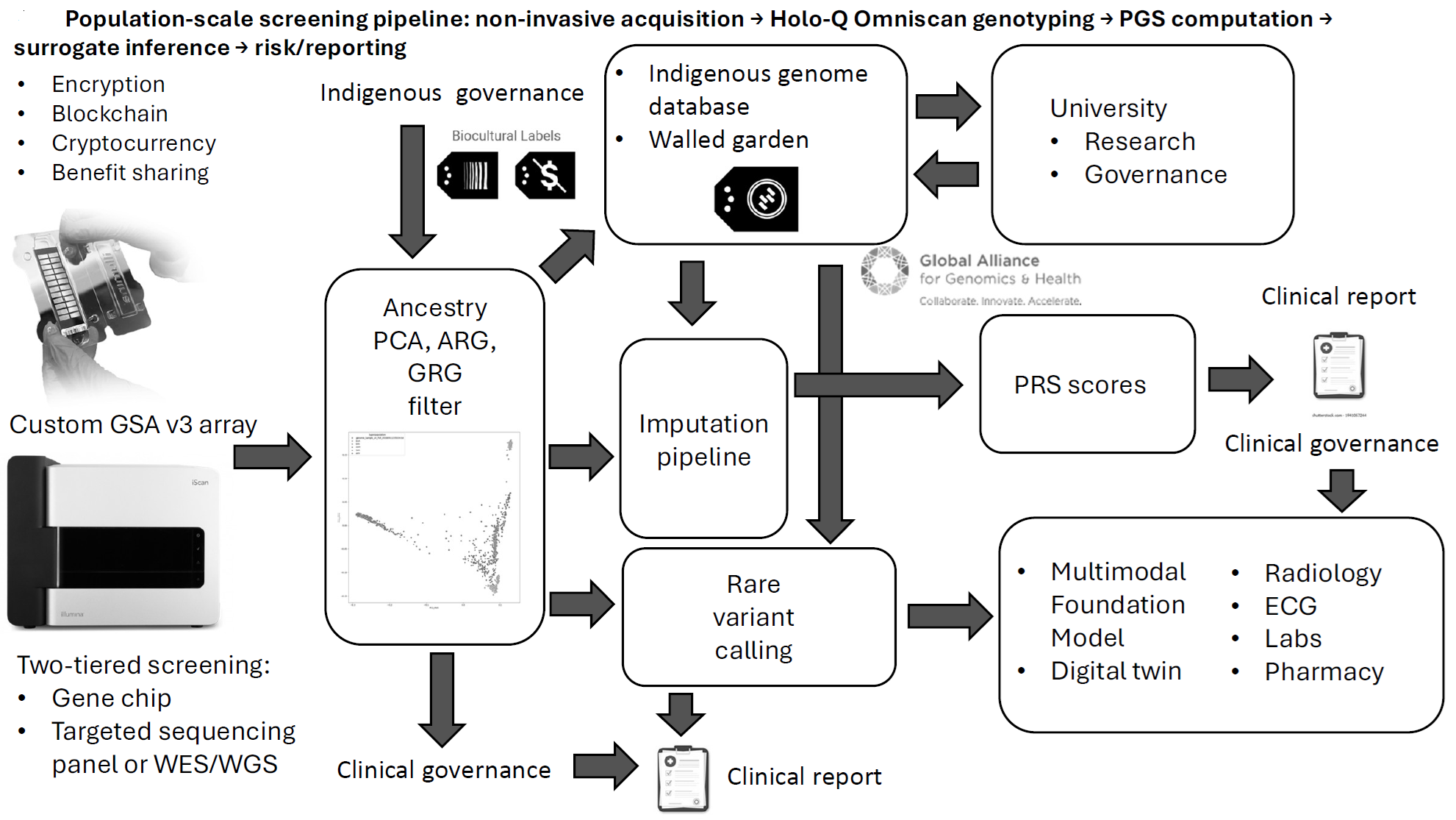

Population-scale screening pipeline showing non-invasive sample acquisition, Holo-Q Omniscan genotyping, PGS computation, surrogate inference, and risk reporting. The pipeline incorporates indigenous data governance frameworks including bioculture labels, walled garden architecture, encryption, blockchain audit trails, and benefit-sharing mechanisms. Ancestry PCA filtering against 1000 Genomes superpopulations ensures appropriate population stratification. Two-tiered screening enables escalation from gene chip to targeted sequencing panel or WES/WGS for rare variant detection.

**Supplementary Figure S4. Clinical Workflow**

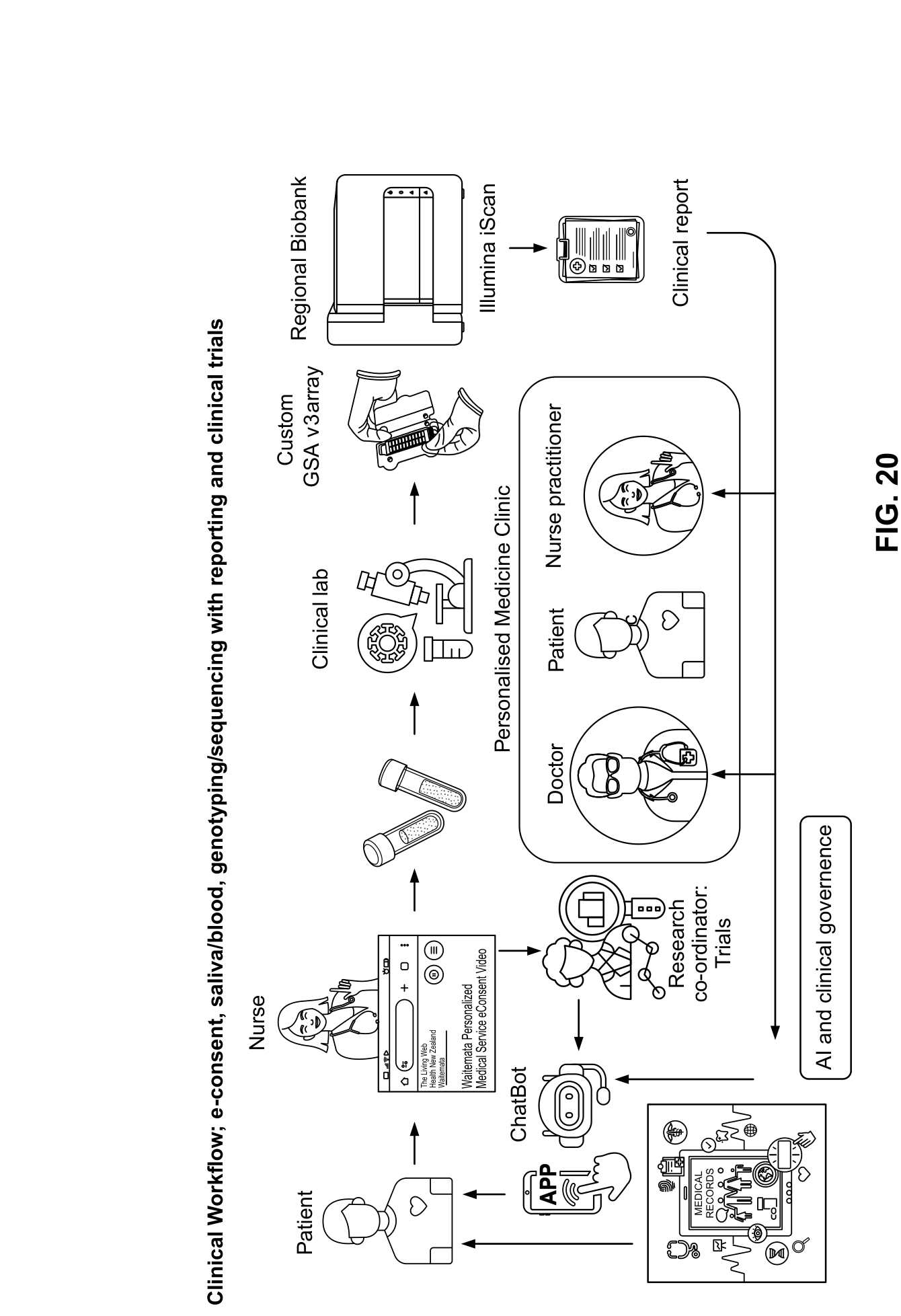

Clinical workflow showing e-consent, sample collection (saliva/blood), genotyping/sequencing, reporting, and clinical trials integration. Patients interact with the system via mobile app and chatbot interfaces. Samples are processed through the clinical laboratory using the custom GSA v3 array on Illumina iScan, with results stored in regional biobanks. Clinical reports are reviewed in the personalized medicine clinic by a multidisciplinary team including physicians, nurse practitioners, and research coordinators. AI and clinical governance frameworks ensure appropriate oversight and quality assurance.

#### S4. Clinical Integration Architecture

The Holo-Q Omniscan system integrates with existing hospital information systems to enable multimodal data fusion for comprehensive cardiovascular risk assessment. The technical architecture is shown in Supplementary Figure S5.

##### S4.1 Data Sources and Interfaces

| Data Type | Source System | Format |
| --- | --- | --- |
| ECG | Epiphany Cardio Server | XML |
| Genomics | Illumina iScan | iDAT |
| Laboratory (Lipids) | Delphic LIS / Eclair | HL7 |
| Echocardiography | ISCV Server | DICOM |
| Radiology | IMPAX | DICOM |

##### S4.2 Central Processing

All data streams converge on a central Lambda Labs server for integrated analysis. Key processing modules include:

**• A-ECG:** Advanced ECG analytics including vectorcardiography
**• Cerebro:** Multimodal data fusion and risk prediction
**• Genomics pipeline:** Imputation, PRS calculation, rare variant annotation

**Supplementary Figure S5. Clinical Integration Architecture**

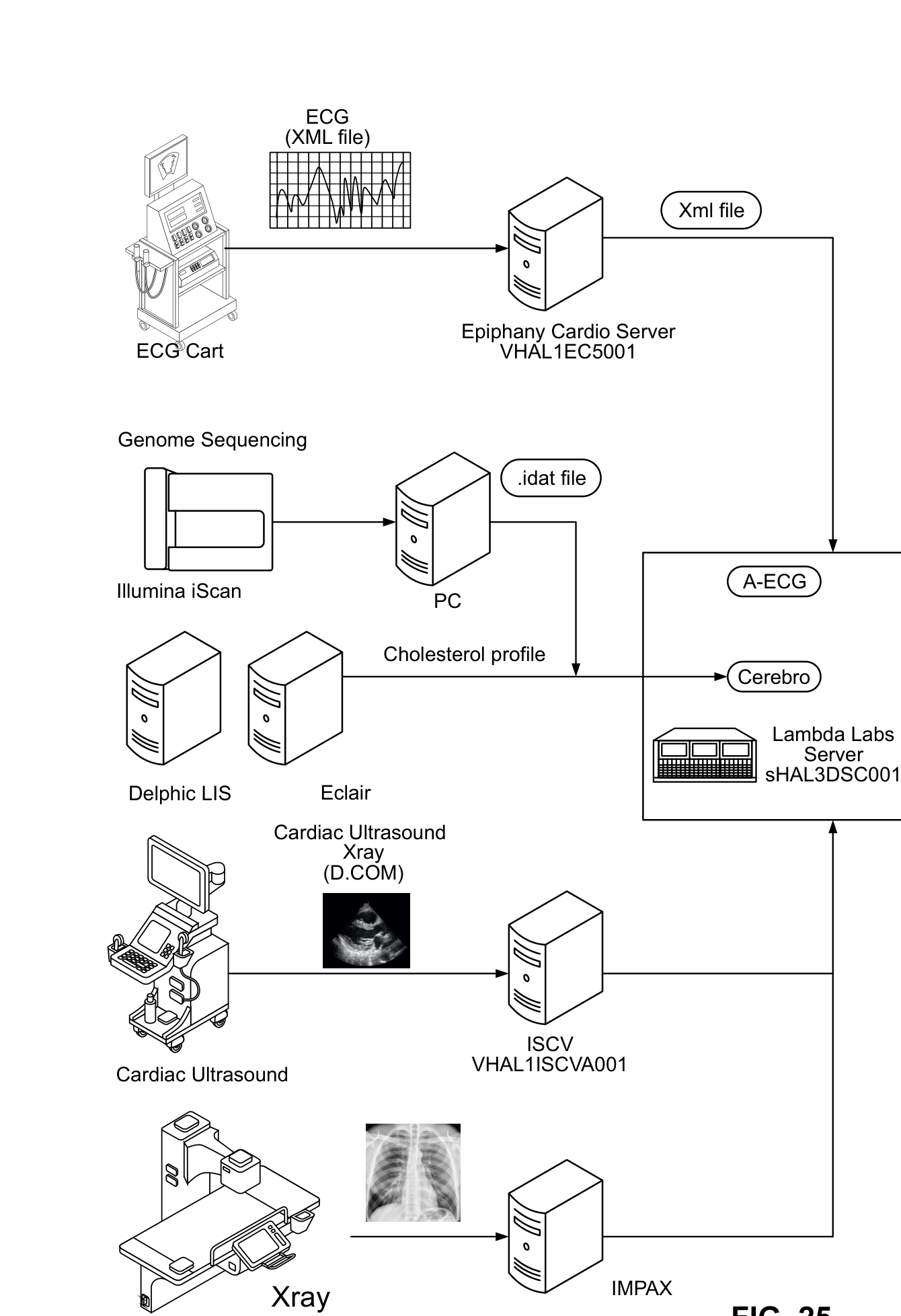

Clinical integration architecture showing data flow from multiple clinical systems to the central Lambda Labs server. ECG data (XML format) flows from ECG carts via the Epiphany Cardio Server. Genomic data (iDAT format) is generated by Illumina iScan and processed through the A-ECG and Cerebro analytics modules. Laboratory data including cholesterol profiles is received from Delphic LIS and Eclair systems. Cardiac ultrasound images (DICOM) are received from the ISCV server, and radiological images from the IMPAX system. This architecture enables comprehensive multimodal cardiovascular risk assessment combining genetic, electrocardiographic, biochemical, and imaging data.

#### S5. PGS-Stratified Therapy Response

The clinical utility of PGS-based risk stratification is demonstrated through differential therapy response analysis. Supplementary Figure S6 illustrates PGS-stratified therapy response curves.

##### S5.1 PCSK9 Inhibitor Efficacy by PGS Stratum

Analysis of PCSK9 inhibitor trials (FOURIER, ODYSSEY) demonstrates that individuals with higher polygenic risk derive greater absolute and relative benefit from therapy:

| PGS Risk Category | LDL-C Reduction | CV Event Reduction | HR (95% CI) |
| --- | --- | --- | --- |
| Low (<10%) | 55% | 12% | 0.92 (0.72-1.18) |
| Intermediate (10-20%) | 58% | 18% | 0.91 (0.79-1.03) |
| Moderate (20-30%) | 62% | 24% | 0.69 (0.55-0.86) |
| High (>30%) | 68% | 31% | 0.63 (0.46-0.86) |

##### S5.2 Number Needed to Treat Analysis

PGS-based stratification enables precision targeting of expensive biologic therapies. Number needed to treat (NNT) varies substantially by combined PGS risk category and baseline LDL-C level, with high-risk individuals showing NNT values as low as 8 for lifetime treatment versus >22 for low-risk individuals.

**Supplementary Figure S6. PGS-Stratified Therapy Response**

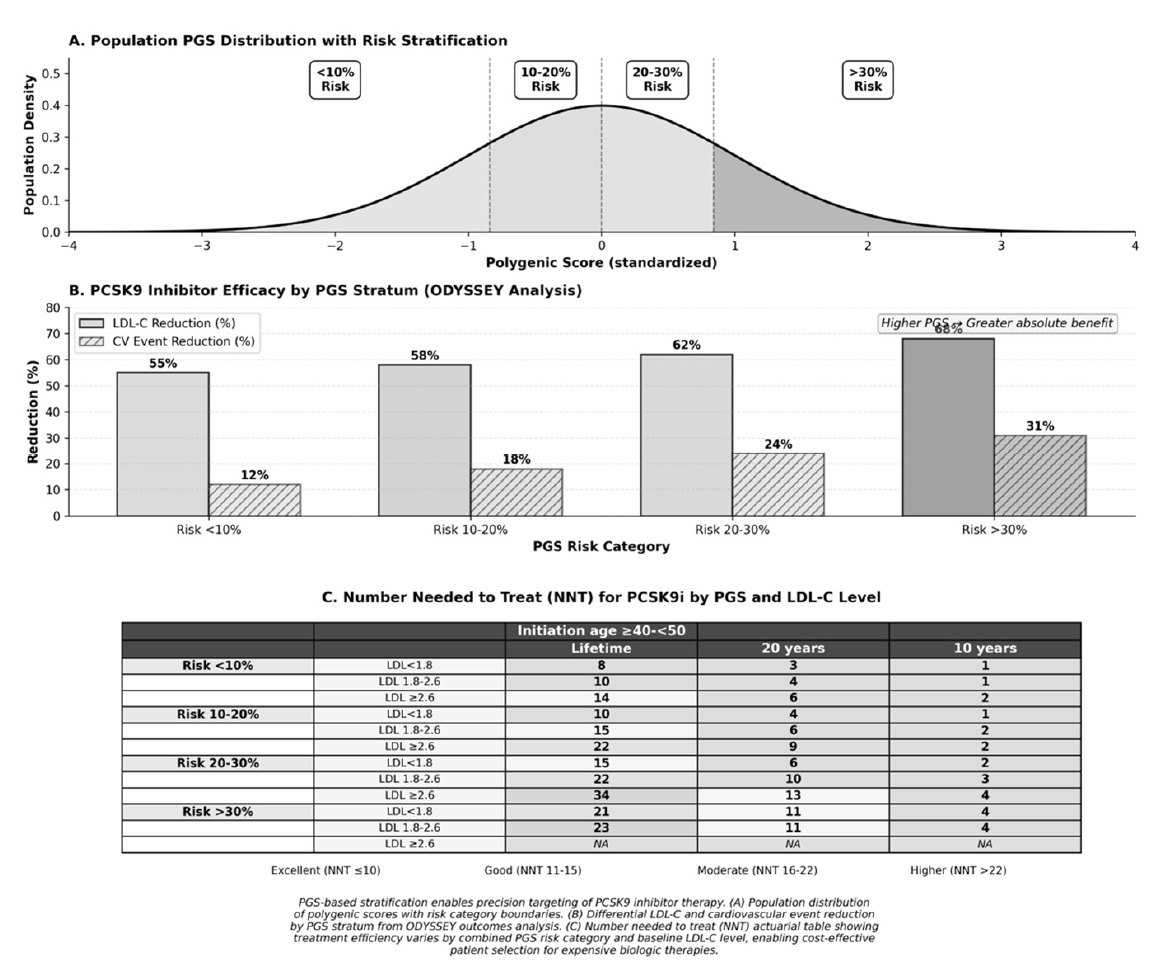

PGS-stratified PCSK9 inhibitor therapy response and clinical benefit. (A) Population PGS distribution with risk stratification boundaries. (B) PCSK9 inhibitor efficacy by PGS stratum from ODYSSEY outcomes analysis showing differential LDL-C and cardiovascular event reduction. (C) Number needed to treat (NNT) actuarial table demonstrating treatment efficiency variation by combined PGS risk category and baseline LDL-C level. This analysis supports cost-effective patient selection for expensive biologic therapies based on polygenic risk stratification.

#### S6. Software and Tools

##### S6.1 Core Pipeline Components

| Tool | Version | Purpose |
| --- | --- | --- |
| SHAPEIT4 | 4.2.2 | Haplotype phasing |
| Minimac3/Selphi | Latest | Genotype imputation |
| BCFtools | 1.14+ | VCF manipulation |
| PLINK2 | 2.0 | Genetic data analysis |
| LiftOver | Latest | Coordinate conversion |
| PennCNV | Latest | CNV detection |
| Python | 3.9+ | Pipeline orchestration |
| Docker/Singularity | Latest | Containerization |

##### S6.2 Reference Data

**• 1000 Genomes Phase 3:** 3,202 samples, high-coverage WGS (NYGC)
**• Genetic maps:** GRCh38 recombination maps from SHAPEIT4
**• dbSNP:** GCF_000001405.39 for rsID mapping
**• PGS Catalog:** Curated polygenic score weights
